# Molecular epidemiology of carbapenem-resistant *Acinetobacter baumannii* strains in Belgian acute-care hospitals

**DOI:** 10.1101/2021.07.06.21259728

**Authors:** Adam Valcek, Pierre Bogaerts, Olivier Denis, Te-Din Huang, Charles Van der Henst

## Abstract

**Objectives:** To describe the genotypic epidemiological distribution and the antibiotic resistance mechanisms of recent carbapenem-resistant *Acinetobacter baumannii* (CRAb) strains recovered from clinical samples in Belgium.

**Methods:** A total of 40 clinical isolates of CRAb collected by the national reference center from 19 acute-care hospitals through national microbiological surveillance in 2014 and 2017 were analysed in this study. The isolates were tested for antimicrobial susceptibility by broth microdilution and determined for carbapenemase-encoding genes by multiplex PCR targeting major carbapenemases families. Isolates were subjected to whole-genome sequencing (WGS) with Illumina technology and the complete chromosomal sequences were *de novo* assembled. Genome analysis was performed to identify intrinsic and acquired resistance determinants and to characterize clonal lineage according to the sequence type (ST).

**Results:** All 40 isolates were resistant to carbapenems and exhibited extensively drug-resistant phenotype with *bla*_OXA-23_ (n=29) being the most abundant detected acquired AMR gene with 38 isolates encoding at least two different types of OXA enzymes. The majority of the isolates were globally disseminated clones of ST2 (n=25) while less frequent sequence types such as ST636 (n=6), ST1 (n=3), ST85 (n=2) and per one isolate from ST604, ST215, ST158 and ST78 were also detected.

**Conclusions:** We have detected extensively drug-resistant globally occurring clones of *A. baumannii* ST1 and ST2 throughout Belgium as well as other sporadic ST including ST636 causing local outbreaks. Our results show the presence of high-risk clones of *A. baumannii* with common travel importation and the crucial need of constant surveillance.

## Introduction

*Acinetobacter baumannii* is a Gram-negative opportunistic pathogen, now recognized as a major hospital pathogen often resistant to multiple major antimicrobials, along with the ability to survive desiccation and disinfectants (1). The emergence of multidrug-resistant *A. baumannii* was introduced to the list of priority pathogens resistant to antibiotics by both WHO and CDC, as it has become a major global threat. *A. baumannii* is capable of acquiring the resistance to different clinically relevant antibiotics such as carbapenems and cephalosporins, limiting the therapeutic options and therefore leading to treatment failure (2). Carbapenemase-resistant *A. baumannii* (CRAb) isolates have also shown increased global spread with rates reaching or exceeding 90% in some clinical settings in Southern and Eastern European countries and elsewhere (3), and mortality rates for the most common CRAb infections such as bloodstream infections and hospital acquired pneumoniae approaching 60% (4). Most of CRAb isolates harbored genes encoding acquired carbapenem hydrolyzing class D β-lactamases and/or class B metallo-β-lactamases. Some CRAb isolates produce an intrinsic OXA-51-like carbapenemase, whereas others produce class B metallo-β-lactamases, including IMP and NDM types (5). The OXA-type carbapenemases represent the most prevalent mechanism of carbapenem resistance in this species, with OXA-23, -24, and -58 being the most prevalent (6). Between 2009 and 2011, OXA-23 producers emerged and replaced the previously predominant OXA-58 producing *A. baumannii* strains (7). The last-line therapeutic options for CRAb isolates are often limited and rely mainly on colistin. Cefiderocol is a novel siderophore cephalosporin with a broad spectrum of activity against Gram-negative pathogens including against MDR *A. baumannii* possessing OXA carbapenemases and/or extended-spectrum beta-lactamases (ESBLs) (8).

In this study, we sequenced the whole genome of 40 clinical isolates of CRAb obtained from Belgian acute-care hospitals and evaluated their resistance phenotype and genotype to describe the recent epidemiological distribution of CRAb in Belgium.

## Materials and Methods

### Bacterial isolates and antimicrobial susceptibility testing

A total of 40 clinical *A. baumannii* isolates (Table 1) were collected from variable samples in patients hospitalized in 19 different hospitals (designated with letters A-S) during two open national passive surveillance surveys in 2014 (n=9) and in 2017 (n=31) for their resistance to meropenem based on local antimicrobial susceptibility testing (AST) methods performed by clinical microbiology laboratories of the hospitals. These isolates were referred on voluntary basis to the National Reference Center (NRC) Laboratory for Antibiotic-Resistant Gram-Negative Bacilli (CHU UCL Namur, Yvoir, Belgium) for confirmation and characterization of carbapenem resistance mechanisms.

**Table 1:**
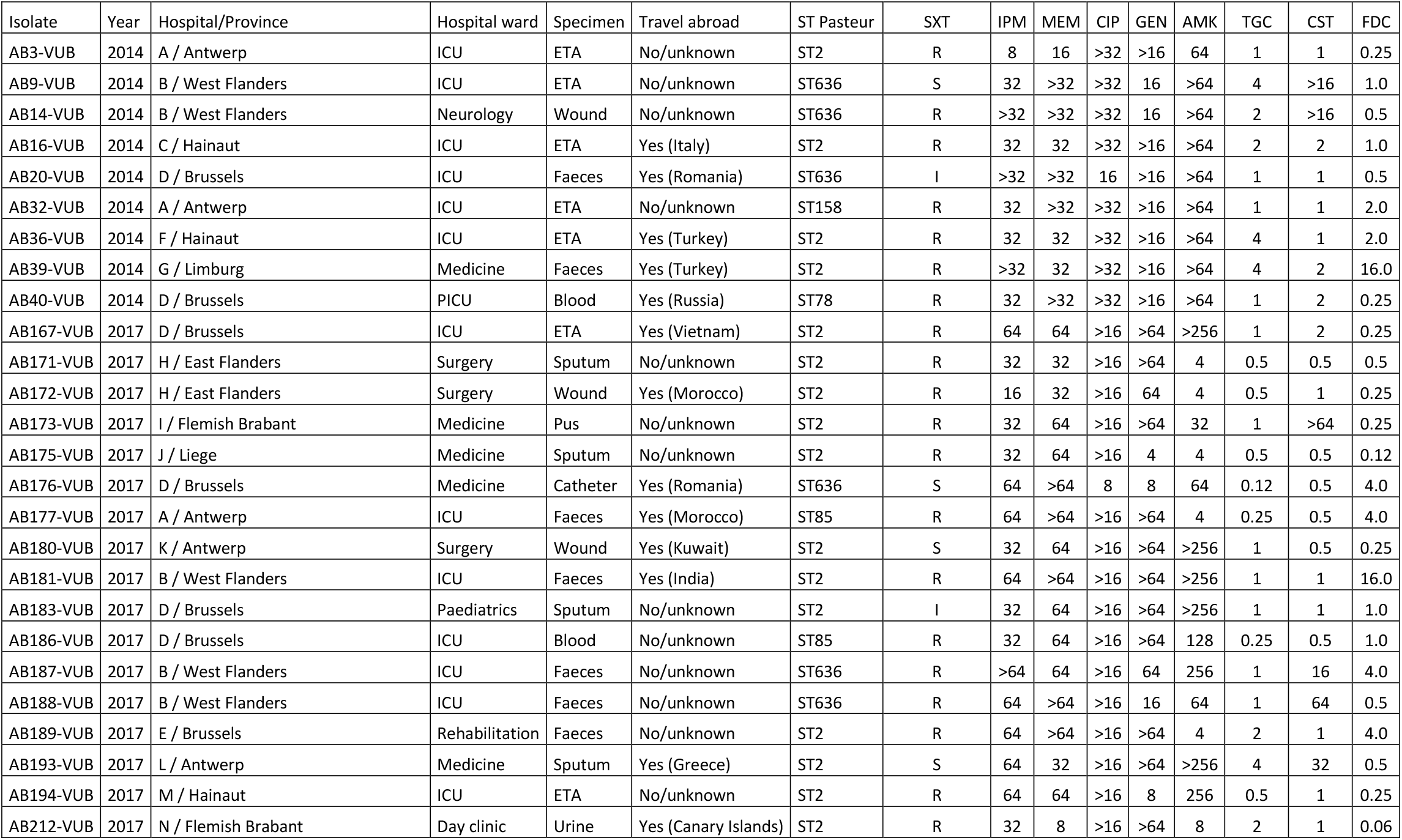
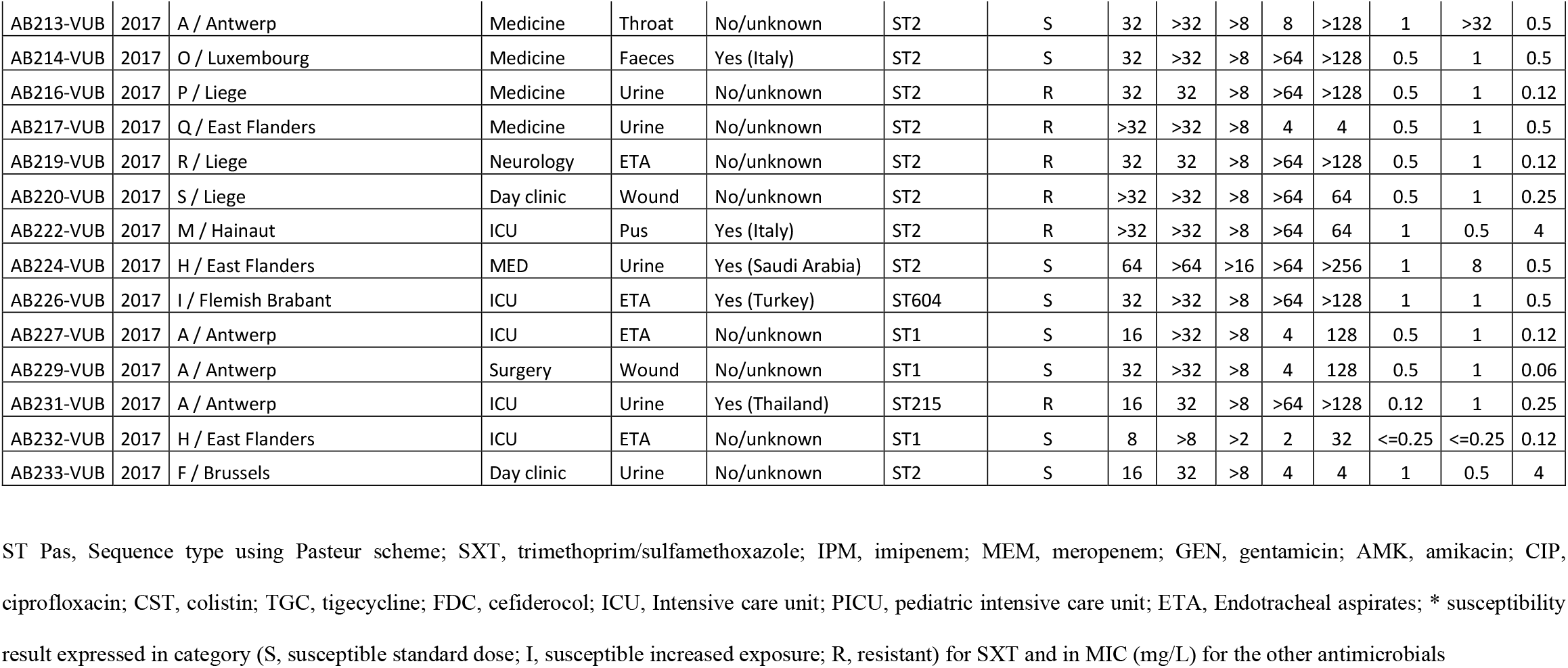
Characteristics and susceptibility results of *A. baumannii* clinical isolates

For the study at the NRC, all isolates were confirmed as *A. baumannii* by MALDI-TOF mass spectrometry (MALDI Biotyper, Bruler Daltonics). AST was performed by paper disk-diffusion (BioRad) for trimethoprim/sulfamethoxazole (SXT) and MICs were determined by broth microdilution (BMD) method using Sensititre customized panels (BEGN5, Thermofisher Scientific) and interpreted according to the EUCAST 2021 clinical breakpoints (Breakpoint tables for interpretation of MICs and zone diameters, Version 11.0, available at https://eucast.org/clinical_breakpoints/) for imipenem (IPM), meropenem (MEM), gentamicin (GEN), amikacin (AMK), ciprofloxacin (CIP), and colistin (CST). MIC obtained for tigecycline (TGC) and cefiderocol (FDC) were interpreted using PK/PD breakpoints (EUCAST 2021, v11.0).

### Whole-genome sequencing and data analysis

A total of 40 clinical isolates were subjected to whole-genome sequencing (WGS) using short reads (Illumina) and *de novo* assembly of the draft genomes. The sub-cultured isolates for AST were used for DNA extraction and following independent sequencing and bioinformatical analyses. Seeing the high genomic dynamics of *A. baumannii* bacteria, we followed the nomenclature in the field (9) by renaming the sub-cultured strains by adding “-VUB”, although these strains are *a priori* identical or very similar.

The genomic DNA was extracted using the phenol/chloroform method. Stationary phase bacteria (2 ml) at an OD_600nm_ of 4 were centrifugated for 1 min at 12.000 x g and resuspended in 200 µl of breaking buffer (2% Triton X-100, 1% SDS, 100 mM NaCl, 10 mM Tris, pH 8.0, 1 mM EDTA, pH 8.0). Then, a ∼200 µl volume of glass beads, and 200 µl of phenol/chloroform were added and vortex at low speed for 30 seconds. 200 µl of TE buffer (10 mM Tris and 1 mM EDTA, pH 8.0) were added, mixed and centrifugated for 5’ at 7.000 x g. The supernatant was transferred into a new Eppendorf tube and 400 µl of phenol/chloroform were added. After centrifugation, the aqueous layer was transferred to a new recipient tube and 1 ml of 100% ethanol was added, mixed and centrifuged for 3’ at 12.000 x g. The supernatant was then removed, the pellet resuspended with 400 µl of TE buffer and 30 µl of 1 mg/ml RNase was added. After incubation for 15’ at 37°C, 10 µl of 4 M of ammonium acetate were mixed, then 1 ml of ethanol 100% was added. After centrifugation (5’ at 12.000 x g), the pellet was resuspended in 100 µl of TE buffer and the final DNA concentration determined by spectrophotometry.

The sequencing libraries were prepared using Nextera XT and subjected to 2×250bp paired-end sequencing on MiSeq (Illumina) using V3_600 kit. The fastq files were generated and demultiplexed using bcl2fastq (Illumina). The sequencing reads were trimmed for quality (Q≤20) and the adaptor residues using Trimmomatic v0.36 (10). The *de novo* assembly was performed using SPAdes v3.15.2 (11) with --careful option.

### Genotypic characterization

The complete chromosomal sequences were subjected to multi-locus sequence typing (MLST) using mlst (https://github.com/tseemann/mlst) employing PubMLST database (12) based on the Pasteur scheme. The resistance genes were detected using Abricate (https://github.com/tseemann/abricate) employing ResFinder (13) with 95% threshold for both identity and coverage. The point mutations were characterized using BLAST algorithm and Geneious R9 (Biomatters, New Zealand).

### Phylogenetic analysis

The maximum-likelihood tree depicting the relatedness of the isolates was constructed from assembled complete genomes using precited open reading frames obtained by Prokka (14) as an input for the core-genome alignment created using Roary (15). RAxML (16) was used for calculation of the phylogenetic tree using general time reversible with optimization of substitution rates under GAMMA model of rate heterogeneity method supported by 500 bootstraps. The phylogenetic tree was visualized and completed with metadata in iTOL (17).

## Results and Discussion

In our study, the 40 clinical isolates exhibited extensively drug-resistant phenotype (Table 1) supported by their genotype (Figure 1). All 40 isolates exhibited extensively drug-resistant phenotypes with *bla*_OXA-23_ (n=29) being the most frequent detected acquired beta-lactamase gene, while the other acquired *bla* carbapenemase-encoding genes were *bla*_OXA-58_ (n=2) *bla*_OXA-72_ (n=9) and *bla*_NDM-1_ (n=1). Intrinsic oxacillinases *bla*_OXA-66, -69, 71, -72, -82, -90, -94, -343, -508_ were detected, showing a great variability of clinical isolates of *A. baumannii* in our collection. While the majority (28/29) of *bla*_OXA-23_ were within Tn*2007* which consists of *bla*_OXA-23_ and IS*Aba4*, common in European isolates of *A. baumannii* (18), AB36-VUB carried *bla*_OXA-23_ within Tn*2008* harbouring one copy of IS*Aba1*, which is a major vehicle of *bla*_OXA-23_ in *A. baumannii* isolates from China (19). Both AB212-VUB and AB3-VUB carried *bla*_OXA-58_ within a composite transposon of IS*Pssp2* (IS1 family), while in AB212-VUB the transposon was preceded by IS*Aba125*. Six out of nine *bla*_OXA-72_ genes were within a composite transposon of IS*Aba31*, while *bla*_OXA-72_ of AB20-VUB was flanked by only one copy of IS*Aba31*. We did not detect any mobile genetic element associated with *bla*_OXA-72_ in AB40-VUB and AB177-VUB. The gene *bla*_NDM-1_ carried by isolate AB177-VUB from a patient with travel history to Morocco was embedded within truncated isoform of Tn*125* transposon consisting of IS*Aba125* truncated by IS*Aba14* as detected in NDM-1 producing *A. baumannii* of ST85 from Tunisia (20). We have also detected high prevalence of genes encoding resistance to aminoglycosides [*aac(3)-Ia* (n=14), *aph(3’’)-Ib* (n=26), *aph(3’)-Ia* (n=25)], streptomycin [*aph(6)-Id* (n=26)], sulfonamides [*sul1* (n=22) and *sul2* (n=11)] and tetracycline [*tet*(B) (n=24)] but also to different types of antimicrobials (Figure 1).

**Figure 1:**
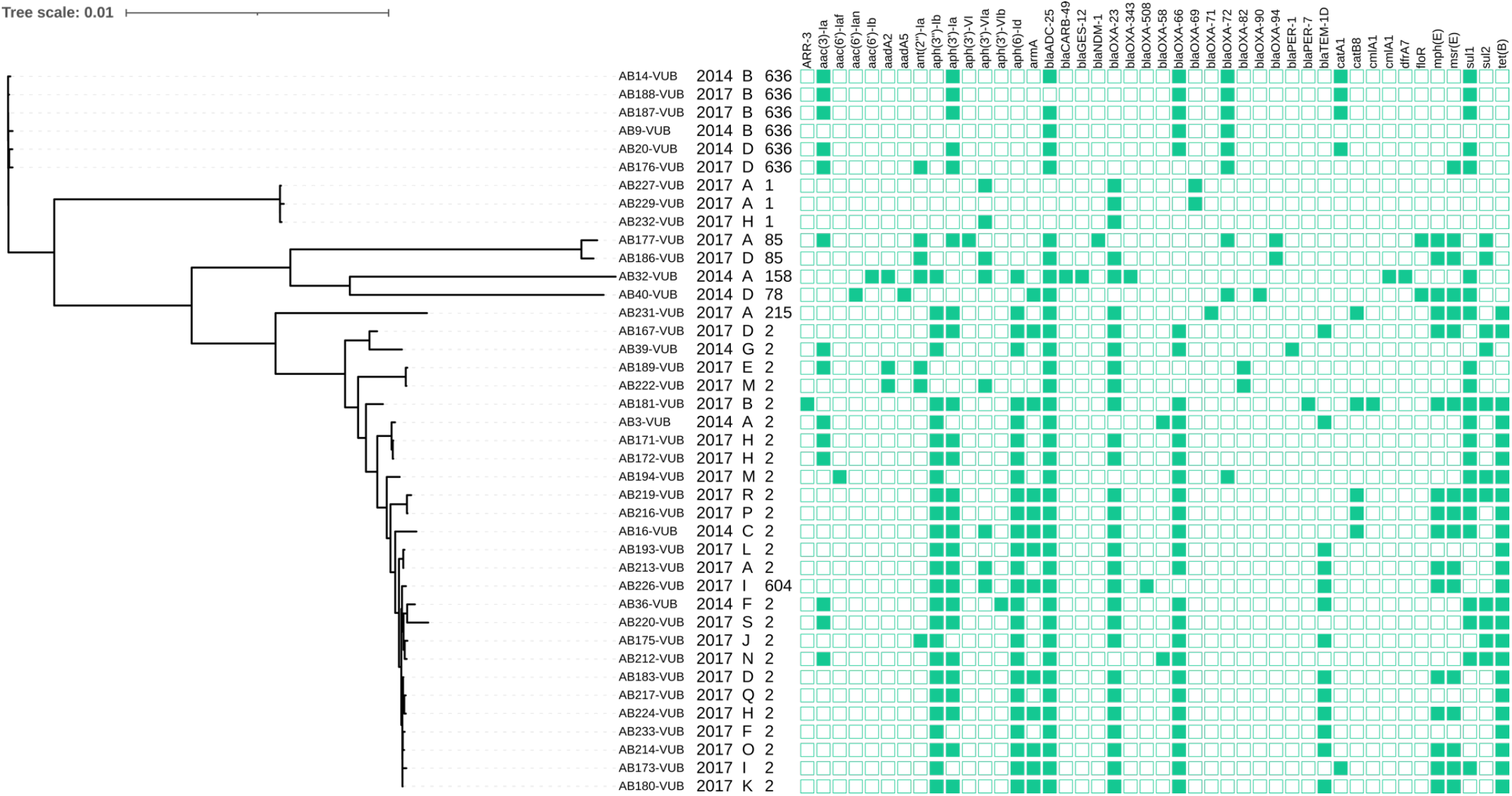
A phylogenetic tree of 40 clinical isolates of *A. baumannii* with depiction of the year of isolation, sampling site (A-S), ST and resistance genes, respectively.

We identified 8 different sequence types (ST), for which ST2 was predominant (n=25), followed by ST636 (n=6), ST1 (n=3), ST85 (n=2) and one isolate from ST604, ST215, ST158 and ST78 (Figure 1). The ST2 and ST1 isolates, previously described as clinically relevant groups, were among the most widely disseminated ST in the complete and draft genomes currently available in the databases. *A. baumannii* ST2 and ST1 account for 71% of all genomes sequenced (21). Isolates of the predominant ST2 were widely distributed in Belgium and carried a broad variety of acquired antimicrobial resistance (AMR) genes including mainly *aph(3’’)-Ib* (n=23), *aph(6)-Id* (n=23), *bla*_OXA-23_ (n=22) and *tet*(B) (n=22). The similar AMR profile and phylogenetic relatedness to ST2 group were detected in AB226-VUB (Figure 1) which belongs to extremely rare ST604 first identified in Egypt (22). Isolates belonging to other rare ST represented per one isolate were of ST215 (AB231-VUB), ST158 (AB32-VUB) and ST78 (AB40-VUB). Two isolates of ST85 (AB177-VUB and AB186-VUB) did not cluster with the major branches of ST2, ST1 or ST636 in the phylogenetic tree (Figure 1), showing their distinct genetic background.

We have detected six isolates of ST636 in two separate hospitals in 2014 and 2017 suggesting the occurrence of clusters. ST636 have been described to cause outbreaks within hospital settings in Serbia and Colombia (23, 24), In our study, two isolates AB176-VUB and AB20-VUB belonging to ST636 detected at one hospital were cultured from two different patients in 2014 and in 2017 with travel history in Romania, indicating possible circulation of *A. baumannii* ST636 in the country.

The *A. baumannii* ST215 is common in Thailand since 2010 (25), which corresponds with the travel history of the colonized patient (Table 1). While GES-producing *A. baumannii* ST158 caused an outbreak in a Tunisian neonatal unit and was linked to GES producing clone from Middle East, but also was identified in Denmark (26), the travel history of the patient infected with AB32-VUB is unknown. The ST78 represented by one isolate (AB40-VUB) was recently detected in Russia as an uncommon clone known as “Italian clone”. Indeed, it has been reported from several Italian hospitals in 2010 and since then it has been detected from other Mediterranean countries, the USA, Germany, Kuwait and French Guiana, pointing towards successful global dissemination (27). Despite ST78 being unusual for Russia, the origin of ST78 (AB40-VUB) in this study is possibly Russian too, as it again corresponds to the travel history of the patient. The ST85 is represented by two isolates (AB177-VUB, AB186-VUB), only AB177-VUB encoded both *bla*_NDM-1_ and *bla*_OXA-94_ and came from a patient with a travel history to Morocco. *A. baumannii* ST85 encoding *bla*_NDM-1_ and *bla*_OXA-94_ genes was previously detected in France, Algeria, Turkey, Syria, Tunisia and recently also Spain (28). The AB186-VUB encoded *bla*_OXA-94_ but not *bla*_NDM-1_ gene, pointing towards geographical unrelatedness of AB177-VUB and AB186-VUB, which is supported by their detection in different hospitals (Figure 1).

Three isolates (AB14-VUB, AB173-VUB and AB188-VUB) exhibited a resistant phenotype to colistin, however none of the isolates harboured *mcr* gene. We have explored the genetic background of these isolates for mutations in the two-component lipid A encoding system *pmrAB*. The isolate AB173-VUB harboured a substitution in *pmrB*^T235I^ while *pmrB*^T235N^ that was described to provide resistance to colistin (29), possibly providing the same colistin-resistant phenotype. Isolates AB14-VUB and AB188-VUB harbored the mutations *pmrB*^L153V^, *pmrB*^N440H^ and *pmrB*^A444V^. However, we identified these mutations in colistin-susceptible isolates too, suggesting a lack of involvement in the resistance mechanism to colistin. We have also excluded interruption of the Lpx pathway as a possible cause of the colistin resistance as the genes *lpxACD* were intact. The mechanism behind the resistance of AB14-VUB and AB188-VUB to colistin remains unclear, however other factors such as outer-membrane asymmetry or efflux pumps might be involved (30). Recent data from 30 European countries showed that 4% of the tested CRAb isolates were resistant to colistin with the majority originating in southern Europe (Greece and Italy) (3).

In our study, cefiderocol retained an activity of 80% overall against the 40 CRAb strains tested showing MIC values ranging 0.12 to 16 mg/l (MIC50 of 0.5 mg/l; MIC90 of 4 mg/l) interpreted according to the PK/PD EUCAST breakpoints (S <=2 mg/l; R >2 mg/l). High MIC of 16 mg/l was detected in two independent isolates belonging to ST2 and expressing both OXA-23 carbapenemase and PER-1 ESBL. Interestingly, our findings are in line with previous studies showing a possible association of PER-type beta-lactamases with higher cefiderocol MICs (31). However other resistance mechanisms to cefiderocol independent from the beta-lactamase production have been proposed including reduced expression of the siderophore receptor by the bacteria (32).

In Belgium, most published data have been related to MDR *A. baumannii* or CRAb outbreaks occurring in single health-care centers following introduction through travel or repatriation abroad from countries outside Belgium before 2010 (33-35). Data from participation to antimicrobial resistance surveillance program showed overall carbapenem resistance among *A. baumannii* ranging 0 to 3.8% during years 2015-2019 (36) and crude incidence density of 0.04 to 0.06 CRAb cases of per 1000 patient-days (37). However, no clear trend could be drawn from these data due to limited number of participation centers or small number of reported isolates and above all, no microbiological characterization and clonal lineage determination were available. The data provided in our study here displayed the first insight in the recent epidemiology of MDR CRAb circulating in Belgian acute-care hospitals and support the current epidemiological status of CRAb spread in Belgium (stage 2b with sporadic hospital outbreaks) according to the latest assessment by the EURGen-Net workgroup (38). Our study also confirmed the continuous importation of CRAb from countries abroad (at least 18 out of the 40 CRAb-carrying patients (45%) had a documented travel history).

Our study demonstrated the wide distribution of internationally disseminated but also travel-imported extensively drug-resistant clones of MDR *A. baumannii* in Belgian health care facilities. These strains pose a serious health issue to the patients especially admitted to high-risk wards such as the intensive care units causing potential nosocomial infections and difficult-to-control outbreaks.

## Data Availability

Data available upon request.

## Data availability

The draft assemblies with the short-read Illumina sequencing reads were deposited in GenBank under BioProject PRJNA734485.

## Acknowledgements

We would like to thank the URBM and URBE research groups from UNamur for the access to their equipment. We also thank Kristina Nesporova for her help in figure elaboration.

## Author contributions

Generation of the *A. baumannii* strain collection, antibiograms, MALDI-TOF mass spectrometry and PCR screening: OD, PB and TDH. Sequencing and bioinformatical analyses: AV and CVDH. Writing of the manuscript: AV, OD, PB, TDH and CVDH.

## Transparency declarations

None to declare.

